# The magnitude and associated factors of low birth weight among newborns delivered in selected public and private hospitals in Addis Ababa, Ethiopia, a comparative cross-sectional study

**DOI:** 10.1101/2025.04.30.25326758

**Authors:** Semere Gebremariam Baraki, Hasset Sisay Asmare, Endale Mengistu Taye

**Affiliations:** Menelik II Medical and Health Science College, Addis Ababa, Ethiopia; Addisu Gebeya Health Center, Gulele Sub-city, Addis Ababa, Ethiopia

**Keywords:** low birth weight, associated factor, public, private, Hospitals, Ethiopia

## Abstract

**Background:** Low Birth Weight (LBW) is one of the primary indicators of the health status of neonates and the nutrition of the mothers. It’s the main determinant of the infant’s survival and growth, both mentally and physically.

**Objectives:** To assess newborns’ low birth weight and its associated factors at public and private health facilities in Addis Ababa, Ethiopia 2023.

**Methods:** An institutional-based comparative cross-sectional study design was employed from April to May 2023. A multistage sampling technique was used to collect data from 533 mothers. The interviewer-administered technique was the method of data collection. Data entry was done by Epi info and analyzed by SPSS. Multivariable logistic regression was employed to identify independent factors to the outcome variable.

**Result:** The overall magnitude of low birth weight was 20.3% (16.9, 23.6, 95% CI). The low birth weight was higher in public hospitals, 24.7% (20.4, 29.8, 95 CI) compared to private hospitals 15.2% (11, 20.1, 95 CI). The neonates born in private and public hospitals have birth weights of means of 3085±681.92 and 2930.21±559.10 grams respectively indicating significant differences (µ1-µ2 = 155.58, 95% CI: (48.60, 262.57) grams. History of abortion (AOR=2.643, 95% CI: 1.309,5.338). History of stillbirth (AOR=3.173, 95% CI:1.158, 8.693), history of chronic disease (AOR= 2.990, 95% CI: 1.414, 6.321), income level (AOR= 5.304, 95% CI: 1.795,15.673), number ANC visit (AOR=2.545, 95% CI: 1.272, 5.091), and gestational age at birth (AOR= .205,95% CI:.095, .441) were found as significant factors that are associated with low birth weight of babies public hospitals, while History of chronic Disease (AOR= 4.056, 95% CI: 1.317,12.492), additional nutrition intake (AOR=4.626, 95% CI: 1.426,15.010) and Gestational age (AOR=.078, 95% CI: .028, .214) were found as significant factors associated with low birth weight of babies born in private hospitals.

**Conclusion and Recommendation:** LBW is a significant public health concern that is linked to multiple factors. The magnitude of LBW and its associated complications would be decreased by the identification of risky mothers and early detection and management of those factors at public and private hospitals.

## INTRODUCTION

World Health Organization defined LBW as the birth weight of a newborn less than 2500 grams at birth weight measurement and remains a major public health problem throughout the world. (1). LBW is a global public health challenge, particularly in sub-Saharan Africa and Asia (2).

More than 20.5 million babies are born with LBW and the magnitude varies by region, which ranges from 7.2% in more developed regions to 17.3% in Asia. (3). Half of all low birth weight babies are born in South and Central Asia, where 27% of babies are born weighing less than 2500 g, while the LBW figures in sub-Saharan Africa are estimated at 15 to 20% (4). According to EDHS 2016, the magnitude of LBW in Ethiopia is estimated at 13%(5). The magnitude of LBW ranged from 6% to 29% in various studies conducted in Ethiopia. (6).

Worldwide, low birth weight is responsible for 60% to 80% of all neonatal deaths, both in developed and developing countries. (7). LBW is an important cause of prenatal mortality and short- and long-term morbidity in infants and children. Deaths of LBW infants are 30 times more common than deaths of normal birth-weight neonates, and they are much more likely to end up with long-term disabilities.(8). The consequences of LBW also cause a higher risk of poor cognition.

Development, and increased risk of chronic diseases later in life prenatal death, and stunting in adulthood, which in turn leads to an intergenerational effect of malnutrition in the affected community and also increases the risk for noncommunicable diseases such as diabetes and cardiovascular disease later in life (9,10).

The majority of LBW newborns in developing countries are due to low gestational age, whereas in developed countries preterm birth is the most common cause of LBW (11). On the other hand, Various physical, and prenatal health statuses, and health follow-ups during pregnancy matter the magnitude of LBW nations. For instance, Low maternal anthropometric measurements, early maternal age, premature parity, low education level, poor maternal nutrition before and during pregnancy, and multiple gestations are also linked to low birth weight(12).

Progress in lowering newborn fatalities has a big impact on development objectives. Since low birth weight is one of the main causes of neonatal mortality, achieving a high coverage of evidence-based interventions that reduce low birth weight and increase the survival of these newborns will be crucial to the achievement of the sustainable development goals (13).

The weight of a newborn at birth is an important indicator of the nutritional status of the mother and the health of the fetus. Worldwide LBW is a major public health problem and a leading cause of neonatal death. Each year, more than 20.5 million newborns are delivered at low birth weight. (14). LBW infants are 20 times more likely to develop complications and die in comparison to normal-weight babies (15).

In Ethiopia, a systematic review study that was done in 2019 shows the magnitude of LBW was 17.3%. The risk factors for LBW are, maternal factors such as poor nutrition, maternal age, inadequate weight gain during pregnancy, malaria, anemia, smoking, alcohol, placental problems, drug use, antihypertensive medications, antidiabetic medications, multiple pregnancies, preterm delivery, intrauterine infection during pregnancy, chronic disease, previous history of LBW and low socio-economic status. (16).

Low birth weight infants are at particular risk of death in their early childhood. Even those who escape death in their first months and years of life are vulnerable to chronic diseases such as diabetes mellitus (II), cardiovascular disease, hypertension, immune deficiency, and impaired language development in adulthood and old age. It also affects reproduction, i.e. premature births and low birth weight, and neurocognitive development such as intellectual disability, learning disability, cerebral palsy, retinopathy, and intellectual disability. (17). In developing countries, low birth weight newborns who survive have poor immune function and increased risk of disease; they are prone to malnutrition, have lower muscle strength throughout their lives, and are more likely to suffer from cardiovascular disease, cognitive disability, and lower IQ, Over the past 10 years, the incidence of LBW in sub-Saharan Africa (SSA) and Asia has not declined. (18).

However, the country has very high neonatal mortality due to factors associated with LBW, which is one of the critical problems that cause short- and long-term health outcomes and mortality in infants. (19).

Despite the World Health Assembly’s target to decrease LBW by 30% by the end of 2025., the progress has stagnated since 2000. To reduce the magnitude of LBW various initiatives implemented targeted in the prenatal, antenatal, intranasal, and postnatal periods. This would result in a relative reduction of 3.9% per year between 2012 and 2025 and a reduction from approximately 20 million to approximately 14 million newborns with LBW at birth. (20). On average, one out of four individuals get maternal and child health care in a private facility. In addition to this, one out of five pregnant women attend a delivery private facility. Out of these, the majority went to a private hospital (68%), a non-government facility (24%), and a private clinic (12%). (21)

Despite tremendous prevention opportunities and efforts, LBW remains a formidable public health challenge in the 21st century, and more detailed studies are needed to explore the factors that influence LBW (22). Although efforts have been made to reduce the incidence of LBW worldwide, LBW is a public health problem globally, in Africa including Ethiopia.(23). Despite all these findings, no evidence indicated the magnitude of LBW and associated factors in private hospitals in Ethiopia. Therefore, this study aimed to describe the magnitude of LBW and associated factors of LBW in both private and public hospitals.

## METHODS AND MATERIALS

### Study Area and Period

Addis Ababa is the capital city of Ethiopia and was established in 1886. The current metro area population of Addis Ababa in 2023 is 5,461,000, a 4.46% increase from 2022. The metro area population of Addis Ababa in 2022 was 5,228,000, a 4.43% increase from 2021. The metro area population of Addis Ababa in 2021 was 5,006,000, a 4.42% increase from 2020 (24). This study was conducted at four public hospitals namely: Yekatit 12 Hospital Medical College (Abebech Gobena MCH), Gandhi Memorial General Hospital, Saint Paul’s Hospital Millennium Medical College, and Tirunesh Beijing Hospital and six private hospitals namely, Ethio Tebib General Hospital, Teklehaimanot General Hospital, Hayat Hospital, Hallelujah General Hospital, TZNA General Hospital, and Amin General Hospital (private hospitals) from April – May 2023.

### Study design

An institutional-based comparative cross-sectional study design was conducted to describe magnitude and identify associated factors of low birth weight, in Addis Ababa from April to May 2023.

### source population

The source population was all newborns delivered to public and private hospitals in Addis Ababa.

### study population

All newborns delivered at selected public and private hospitals of Addis Ababa during the data collection period were the study population.

### Eligibility criteria

#### Inclusion criteria

All newborns delivered at Addis Ababa Ethiopia, in selected public and private health facilities, were eligible irrespective of their gestational age.

#### Exclusion criteria

- Newborn babies whose mothers suffered from severe medical or surgical health problems
- Twin delivery,
- Newborn delivered with congenital anomalies,
- unknown last normal menstrual period

### Sample Size Determination

The researcher used a double proportion formula by considering the P1= the proportion of low birth weight in public hospitals taken from a study done in Addis Ababa which is 14% (25). And P2= the proportion of low birth weight in private hospitals which is 5 % taken from the survey taken before the actual data collection from private hospitals with a design effect of 1.5 gave the final sample size of = **<u>561</u>**

### Sampling Techniques and Procedure

First, we identified the public and private hospitals that give delivery service in Addis Ababa, then we selected four hospitals from the public and seven hospitals from private institutions. They select all hospitals using simple random sampling. A multi-stage sampling technique was used to invite newborns and include them in the study. Thirty percent of both public and Private General Hospitals (4 public and 7 private general hospitals) were included in the study. Proportional size allocation will be used to select the study population from each selected private and public hospital.

### Variables Dependent Variable

Low Birth Weight

### Independent Variables

**Socio-demographic characteristics**: age of mother, marital status, educational status, occupational status, residence, income level,

**Nutritional characteristics:** iron and foliate supplementation, dietary advice during ANC, additional dietary, maternal MUAC, and maternal, Anemia

**Obstetrics and neonatal characteristics:** parity, birth interval, condition of pregnancy, gestational age at birth, history of family planning, the weight of new birth, sex of newborn, of ANC starting time, ANC visit frequency, History of abortion in the previous pregnancies, History of twin pregnancy, History of stillbirth previous, history of low birth weight and chronic disease.

**Behavioral characteristics:** history of drinking alcohol and chewing chats and cigarette smoking mother’s work condition.

### Operational Definition

**Birth at Privat hospital**: When the mother had a follow-up in private and gave birth in a private hospital

**Birth at Public hospital**: When the mother had a follow-up in private and gave birth in A public hospital

**Chronic disease**: the mother is categorized as having chronic disease if she answer for one of the following chronic disease orally ( hypertension, diabetes, allergy, asthma, kidney problem, or obesity)

**Low birth weight**: was considered, when newborn weight recorded below 2500 g(26)

**Gestational age**: is “the duration of time measured from the first day of conception and expressed in complete weeks” (27).

**Preterm birth**: when a baby is born before 37 weeks of gestational age.

**Public hospitals:** These are governmental hospitals in Addis Ababa City Administration Health Bureau.

**Private hospitals:** hospital not owned by the government founded for profits

### Data Collection tools and quality control

A semi-structured questionnaire and data extraction checklist were adopted by reviewing similar literature and the Ethiopian Demographic Health Survey 2016 (11,22). The questionnaire was designed to measure socio-demographic characteristics, maternal and obstetric, nutritional status characteristics and extra diet and iron supplementation, hemoglobin level, behavioral characteristics, and various factors affecting LBW. The weight of the newborn was measured As soon as possible using standardized weight measurement scale. The principal investigator also prepared data collection in English and then translated into Amharic. Two-day training was conducted for both data collectors and supervisors. Pretest and supervisor were done to ensure the data quality.

### Data Analysis Procedure

The data was entered by Epi-info and analyzed by SPSS (version 25) software. The Bivariate Logistic regression model was used to identify the association between the explanatory (independent) variable and Low birth weight (dependent variables). A p-value <0.2 at bivariate analysis was for a variable to be included in the multivariate analysis. The model fitness was checked using Hosmer and Lemeshow’s goodness of fit (p > 0.05). No multicollinearity was detected among the independent variables. The researchers used Multivariable logistic regression with a p-value of <0.05 identify the predictors of LBW. Adjusted Odd Ratio (AOR) with 95% CI (confidence interval) was used to measure the strength of association between explanatory variables and predicted variable (LBW). An independent t-test was used to test mean differences in birthweight between public and private hospitals and

### Ethical Consideration

The ethical issue letter was written by Menelik II Medical and Health Science College, Research and Community service coordinator. Approved ethical clearance was obtained from the Addis Ababa Public Health Research and Emergency Management directorate. Supportive letters were obtained from colleges to public and private hospitals. The researcher provided detailed information about the research aim and their right to respond fully or partially to the questionnaire, and after assuring all this, we got Informed consent from the study participants. All data was kept confidential; it was used for research purposes only as well and confidentiality was maintained by omitting the name of the respondents.

## RESULT

### Sociodemographic and economic characteristics of study participants

The researcher collected 533 participants with a response rate of 95%. The mean age of study participants was 28.5 ± (5.24). The majority of participants ( 80.3%) were between age of 21-34 years. On the other hand, 45(15.9%) of the participants from the public hospital are government employees while 57 (22.8%) of the participants from the private hospital are governmental employees. Regarding the monthly income, 88 (31.1%) from public hospitals and 131 (51.4%) from private hospitals had monthly income >6001 birr per month (Table 1).

Table 1:

### Obstetric and neonatal-related characteristics

Out of the total respondents, 533 study participants 108(20.3%) had low birth weight, from those, 70(25%) were from public hospitals, and 38(15%) were from private hospitals. There were two hundred.

Seventy-six (51.8%) newborns are males, among the participants155(29.1%) had few ANC< 8 visits from this 97(34.3%) are from public hospitals and 58(23.2%) are from private hospitals, almost 84(15.8%) mothers who had a history of previous low birth weight 21(18%) from public hospitals and 37(14.8%) from private hospitals, more than half 396(74.3%) mothers with LBW babies has less intake of Folic acid before conception, 233 (82.3%)are from public and163(65.2%) private hospitals. Of a substantial number of newborn mothers’ total of 90(16.9%), 68(24%), and private hospitals 22(8.8%) did not intake Iron supplementation during pregnancy. Most of study participants were multigravida 365(66.8%), 195(57.25) public and 173(69.2%) in private hospitals. Mothers with LBW babies who have Chronic disease were 141(26.5%), 65(23%) are from public and private 76(30.4%) hospitals (table 2).

Table 2

### Maternal nutritional and behavioral factors

Out of a total of 283 participants, 47(16.6%) didn’t receive nutritional advice during their pregnancy from public hospitals, whereas out of 250 participants, 31(12.4%) didn’t receive it from private hospitals. Among 283 respondents from public hospitals, 63(22.3%) and 250 mothers from private hospitals 22(8.8%) were underweight. Amongst 283 mothers 40 (14.1%) individuals from public hospitals and out of 250 respondents 29(11.6%) from private hospitals were found to be Anemic. Out of 283 participants 8(2.8%) and out of 250 participants 8 (3.2%) had a history of alcohol usage (Table 3).

Table 3

### Magnitude of LBW in public and private hospitals in Addis Ababa, Ethiopia

Based on the finding’s magnitude of LBW among neonates born in private and public hospitals was 15.2 (11, 20.1) % and 24.7 (20.4, 29.8) respectively. But the total prevalence in all hospitals (public and private) hospitals was 20.3% (95% CI: (16.9, 23.6) (figure 1).

**Figure 1:**
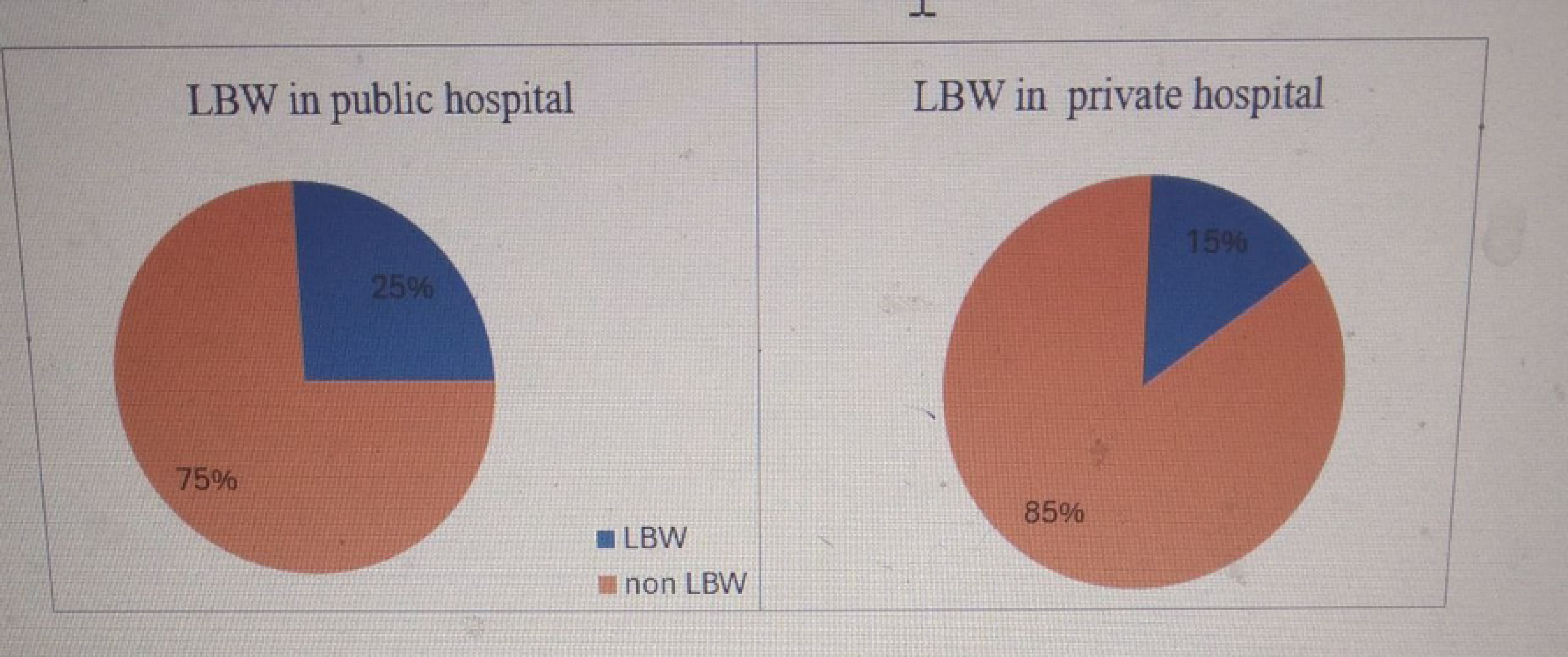
The magnitude of low birth weight in public and private hospitals in Addis Ababa.

The neonates born in private and public hospitals have birth weights of means of 3085±681.92 and 2930.21±559.10 grams respectively. the mean birth weights difference between private, and public has a significant difference which is (µ1-µ2 = 155.58, 95% CI: (48.60, 262.57) grams (Table 4).

Table 4

### Factors associated with low birth weight in selected public hospitals

Based on the bivariate logistic regression, history of family planning, history of abortion, history of low birth weight, history of stillbirth, history of chronic disease, nutritional advice during pregnancy, additional nutrition, income level, hemoglobin level, number of ANC visits, and gestational age were among the variables associated with LBW. However, in the multivariable logistic regression, the history of abortion, history of stillbirth, history of chronic disease, income level, number of ANC visits, and gestational age were significantly associated with LBW in public hospitals. History of abortion (AOR=2.643, 95% CI: 1.309,5.338), history of stillbirth (AOR=3.173, 95% CI:1.158, 8.693), history of chronic disease, (AOR= 2.990, 95% CI: 1.414, 6.321), Income level (AOR= 5.304, 95% CI: 1.795,15.673), number ANC visit (AOR=2.545, 95% CI: 1.272, 5.091), and gestational age at birth (AOR= .205,95% CI:.095, .441) were found as significant factors that are associated with LBW (Table 5)

Table 5

On the other hand, in the bivariable analysis, Employment statutes, History of abortion, History of low birth weight, Iron intake during pregnancy, History of folic intake before pregnancy, History of chronic Disease, Nutritional advice additional nutrition intake, Hemoglobin level, Gestational age were the factors associated with LBW in private hospitals. However, the multivariable logistic regression analysis shows.

History of chronic Disease (AOR= 4.056, 95% CI: 1.317,12.492), Additional nutrition intake (AOR=4.626, 95% CI: 1.426,15.010) and Gestational age (AOR=.078, 95% CI: .028, .214) were found as significant factors that are associated with low birth weight of babies born among mothers attending private hospitals in Addis Ababa, Ethiopia (table 6)

Table 6

## DISCUSSION

This study assessed the newborns’ LBW and associated factors at public and private health hospitals in Addis Ababa, Ethiopia. Overall, the magnitude of low birth weight is 20.3% (16.9, 23.6). The magnitude of low birth weight was higher in public hospitals is 25% (20.4, 29.8) compared to private hospitals is 15 % (11, 20.1).

The finding of this study is various studies done in Ethiopia (19,29,30), but higher than studies conducted in Ethiopia and outside Ethiopia (9,26,31,32). This LBW magnitude and differences may be due to the inclusion of preterm babies in this study. Similarly, the magnitude of low birth weight in this study is higher than in Brazil (33). This difference could be due to socioeconomic conditions, demographic characteristics, organization interventions on maternal health services, including nutrition and women’s economic empowerment, and the accessibility of service delivery could be contributing factors.

Newborns born to mothers who had a history of abortion were 2.643 times more likely to have low birth weight as compared to those who have no history of abortion (AOR=2.643, 95% CI: 1.309, 5.338). This study was in line with the study that was done in India (34). This might be due to that “pregnant women with a history of abortion were more likely to have preeclampsia, premature delivery, and low birth weight in a subsequent pregnancy than those without a history of abortion” (35).

Newborn mothers who had a history of stillbirth were 3.173 times more likely to have low birth weight compared to those who had no history of stillbirth (AOR=3.173, 95% CI: 1.158, 8.693) . Women with a history of stillbirth are at higher risk of other adverse pregnancy outcomes, such as preterm birth, low birth weight, and placental abruption. They may be “at risk for fetal growth restriction in the subsequent pregnancy” (36). Families are uniquely impacted by physical, psychological, and economic problems; and need interventions that should strive to manage the problems sufficiently (36).

Newborns born to mothers who had a history of chronic disease were 2.990 and 4.056 times more likely to have low birth weight as compared to those who have no history of chronic disease in public (AOR=2.990, 95% CI:1.414, 6.321)) and private hospitals (AOR=4.056, 95% CI:1.317,12.492) respectively. This study was supported by the study that was done in Mekelle and Gamogofa. (11, 37). Similarly, the occurrence of chronic conditions among women before pregnancy was associated with a significant burden of adverse pregnancy outcomes such as pre-term and low birth weight. So, preconception intervention could be important in improving maternal and newborn health outcomes. (38)

Newborns born to mothers who had a history of the income level of pregnant mothers (<2000) were more likely to have low birth weight as compared to those who have, an income level of pregnant mothers greater than two thousand (AOR =5.304, 95% CI: 1.795,15.673) in public hospitals. Women belonging to low socio-economic status are at a greater risk of being underweight (39), which eventually leads to giving birth to an LBW child. Few other studies showed that the LBW of a child is determined by the mother’s economic class (40, 41, 42). This may be due to the indirect effect of their income, which enables them to have better access to nutrition and other basic needs before and during pregnancy.

Newborns born to mothers in public hospitals who had less than eight ANC visits were 2.5 times more likely to have LBW than those who had more ANC visits (AOR = 2.545, 95% CI: 1.272, 5.091). Studies done in Uganda, Dilla, and the North Showa zone also support this finding (28, 43,44). This could be because as ANC sessions increase, knowledge of the mother, problem screening by health care providers, and preventive measures can be enhanced more (45). This helps to reduce the odds of giving birth to a child with LBW. So, health professionals should put their efforts towards promoting more ANC attendance.

On the other hand, our study showed that pregnant women attending private hospitals who did not take additional nutrition were 4.6 times more likely to have newborns with LBW as compared to women who took additional nutrition during pregnancy (AOR=4.626, 95% CI:1.426,15.010). This finding was consistent with findings of other studies conducted in West Ethiopia (46), and Ghana (47). Inadequate, nutrition, unbalanced diets, and low dietary diversity might negative impact on birth weight (48), because Undernutrition during pregnancy, especially during the third trimester, is associated with reduced birth weight, which is an indicator of intrauterine growth restriction (49).

Infants born pre-term was found more likely to have LBW compared to term babies in both public and private hospitals. Based on WHO estimations, 25 million babies are annually born with LBW annually (50). Although prematurity is a major reason for LBW, it is not a perfect proxy for preterm birth. Term babies might be born with LBW. It is estimated that estimated there is 67% overlap between preterm birth and LBW (51, 52). “Mechanisms and risk factors for preterm and for LBW babies may differ despite a substantial proportion of LBW being contributed by preterm births as LBW infants are also a result of intrauterine growth restriction” (53)

## CONCLUSION AND RECOMMANDATIONS

The magnitude of LBW in Addis Ababa Ethiopia was found to be 20.3 (16.9, 23.6). It was higher in public hospitals. Risk factors for the low LBW of newborns among mothers attending public hospitals were history of abortion, history of stillbirth), history of chronic disease, income level ANC visits sessions, gestational age at birth, history of chronic disease, additional nutrition, and income level were found risk factors for LBW of newborns among mothers attending private hospitals. LBW is a significant public health concern that is linked to multiple factors. To reduce the magnitude of LBW regarding the aim of national and global targets, additional nutrition advice, increased antenatal care, early diagnosis and treatment of chronic disease should be enhanced more. Previous history of stillbirth and abortion should be properly screened and managed in the current programs by identifying and possible causes of still birth and abortion. Since there is variation in some factors related to LBW in private and public hospitals responsible bodies work to improve

## Strength and Limitation

This study was carried out relatively at different public and private hospitals which enables us to compare the magnitude and associated factors. In this study, however, participant memory and the self-report nature of the determinant factors are possible drawbacks that could introduce bias. The study is cross-sectional, it can indicate the cause-and-effect relationships.

## Ethics statement

The studies involving human participants were reviewed and approved by the Ethical Review Board Menelik II Medical and Health Science College. Written informed consent to participate in this study was provided by the participants’ legal guardian/next of kin.

## Author contributions

SGB and HSA: conceptualization, formal analysis, methodology, resources, software, supervision, validation, visualization, and writing and editing —original draft. EMT: data collection, supervision, and writing and editing of the manuscript. All authors read and approved of the final manuscript.

## Data Availability

The raw data supporting the conclusions of this article will be made available by the authors, without undue reservation

## Acknowledgments

We thank all the respondents and data collectors for their participation in the achievement of this work. We also thank the Menelik II Medical and Health Science College for its ethical clearance.

## LIST OF ABBREVIATIONS AND ACRONYMS

ANC: Ante Natal Care
BMI: Body Mass Index
CSA: Central Statistical Agency
EDHS: Ethiopian Demographic Health Survey
EPY: Ethiopian Fiscal Year
ELBW: Extreme Low Birth Weight
IQ: Intelligent Quotient
LBW: Low Birth Weight
MUAC: Mid Upper Arm Circumference
SSA: Sub-Saharan Africa
UNICEF: United Nations Children’s Fund
VLBW: Very Low Birth Weight
WHO: World Health Organization

